# Perspectives of Women and Healthcare Providers on Facilitators and Barriers to Cervical Cancer Screening in the Rural Kilimanjaro Region Tanzania: A Qualitative Study

**DOI:** 10.1101/2025.08.01.25332778

**Authors:** Rogathe F. Machange, Bernard Njau, Christina C. Mtuya, Victor W. Katiti, Lyidia Masika, Bariki Mchome, Eva Kantelhardt, Mirgissa Kaba, Blandina T. Mmbaga, Declare L. Mushi, Gunilla Björling, Rachel Manongi

**Affiliations:** School of Public Health, KCMC University P.O. Box 2240, Moshi, Kilimanjaro, Tanzania; Kilimanjaro Christian Medical Centre, P.O. Box 3010, Moshi, Tanzania; Kilimanjaro Clinical Research Institute, P.O. Box 2236, Moshi, Kilimanjaro, Tanzania; Martin-Luther-University Halle-Wittenberg, Department of Gynecology, Germany; School of Public Health, Addis Ababa University, Ethiopia; Karolinska Institutet, Department of Neurobiology Care Sciences and Society, Stockholm Sweden; Jönköping University, School of Health and Welfare, Jönköping, Sweden

**Keywords:** facilitators, barriers, cervical cancer screening, Community, TDF, COM-B

## Abstract

Cervical cancer (CC) can be prevented and treated if detected early through regular screening. However, 88% of women aged 30 to 49 have never been screened in Tanzania. This study explored the facilitators and barriers to CC screening among women and healthcare providers in rural Kilimanjaro, Tanzania. We used the Theoretical Domains Framework (TDF) and the Capability, Opportunity, Motivation-Behavior (COM-B) model to identify, code, and synthesize behavioral facilitators and barriers to screening. An exploratory study design was employed, and data were collected from 13^th^ March to 1^st^ October 2024 in rural Kilimanjaro’s districts. Purposive sampling was employed. Six focus group discussions (FGD) were conducted with 54 women aged 30–50 years, including two groups of screened women, two with non-screened women, two groups combining screened and non-screened women, and twelve in-depth individual interviews with healthcare providers. Data were transcribed, coded, and thematically analyzed using QDA Miner Lite v2.0.8. Four main themes emerged: knowledge about CC and screening, the power of social influence, emotional and structural influences on CC screening, and enhancing CC screening uptake. The findings highlighted several barriers to CC screening, including limited knowledge, misconceptions, fear of pain and positive results, limited access to services, and a shortage of trained providers. Despite these challenges, social support from family, peers, community leaders, and healthcare providers’ educational efforts (physical and social opportunity) was identified as a strong facilitator. In conclusion, increasing CC screening uptake in rural communities requires targeted, theory-informed interventions incorporating education that enhance awareness, community engagement, strengthen social support, and improve health services.

## Introduction

Cervical cancer is one of the most preventable and curable forms of cancer, especially when detected and treated early. Despite this, it remains a major public health concern and is the fourth most common cancer among women globally(1). Each year, approximately 660,000 new cases and 350,000 deaths occur, with 94% of deaths happening in low- and middle-income countries. In sub-Saharan Africa, cervical cancer (CC) is the second most common cancer-causing death in women (2). In Tanzania, cervical cancer is the leading cancer among women aged 15–44, with an incidence rate of 9,770 and a mortality rate of 6,695 per 100,000 women (3).

The World Health Organization (WHO) launched a global strategy in 2020 to eliminate CC. The strategy includes screening 70% of women aged 30–49 at least twice in their lifetime and ensuring 90% of women with precancerous lesions receive treatment (3,4). However, Tanzania still faces significant challenges in achieving this target. Despite offering free screening services since 2004 (3), only 12% of eligible women had been screened by 2021–2022, highlighting significant limitations in the national screening program implementation. To increase screening uptake, it’s crucial to understand facilitators and address barriers to CC screening(5).

Early screening is a crucial strategy for preventing CC (6). However, uptake of CC screening among women is low partly due to factors like limited knowledge, fear, privacy concerns, and cultural barriers (7,8). Facilitators such as supportive attitudes, education, and trusted environments have been shown to increase CC screening uptake (9–12). Theory-based interventions, particularly those guided by the Capability, Opportunity, Motivation-Behavior (COM-B) model and Theoretical Domain Framework (TDF), have proven more effective in overcoming barriers and guiding behavior change strategies (13–15). The TDF integrates 33 behavioral theories into 14 domains to identify cognitive, social, and environmental influences on behavior (14,16) *as Table 1*. The COM-B model explains behavior as the result of the interaction between capability, opportunity, and motivation(17) (*see Figure 1*). These frameworks help tailor interventions to the community (*see Table 2)*.

**Table 1:** The Theoretical Domains Framework

| **TDF Domain Knowledge** | **Definition** |
| --- | --- |
| Knowledge | Information relating to the behavior |
| Skills | An ability or proficiency acquired through practice |
| Social/Professional Role and Identity | A coherent set of behaviors and displayed personal qualities of an individual in a social or work setting |
| Beliefs about Capabilities | Acceptance of the truth, reality or validity about an ability, talent or facility |
| Optimism | The confidence that things will happen for the best or that desired goals will be attained |
| Beliefs about Consequences | Acceptance of the truth, reality or validity about outcomes of a behavior in a given situation |
| Reinforcement | Increasing the probability of a response by arranging a dependent relationship, or contingency, between the response and a given stimulus |
| Intentions | A conscious decision to perform a behavior or a resolve to act in a certain way |
| Goals | Mental representations of outcomes or end states that an individual wants to achieve |
| Memory, Attention and Decision Processes | The ability to retain information, focus selectively on aspects of the environment and choose between two or more alternatives |
| Environmental Context and Resources | Any circumstance of a person's situation or environment that discourages or encourages the development of skills and abilities, independence, social competence and adaptive behavior |
| Social influences | Those interpersonal processes that can cause individuals to change their thoughts, feelings or behaviors |
| Emotion | A complex reaction pattern, involving experiential, behavioral, and physiological elements, by which the individual attempts to deal with a personally significant matter or event |
| Behavioral Regulation | Anything aimed at managing or changing objectively observed or measured actions |

Note: Adapted from Cane et al (2012) <sup>16</sup>

**Figure 1:**
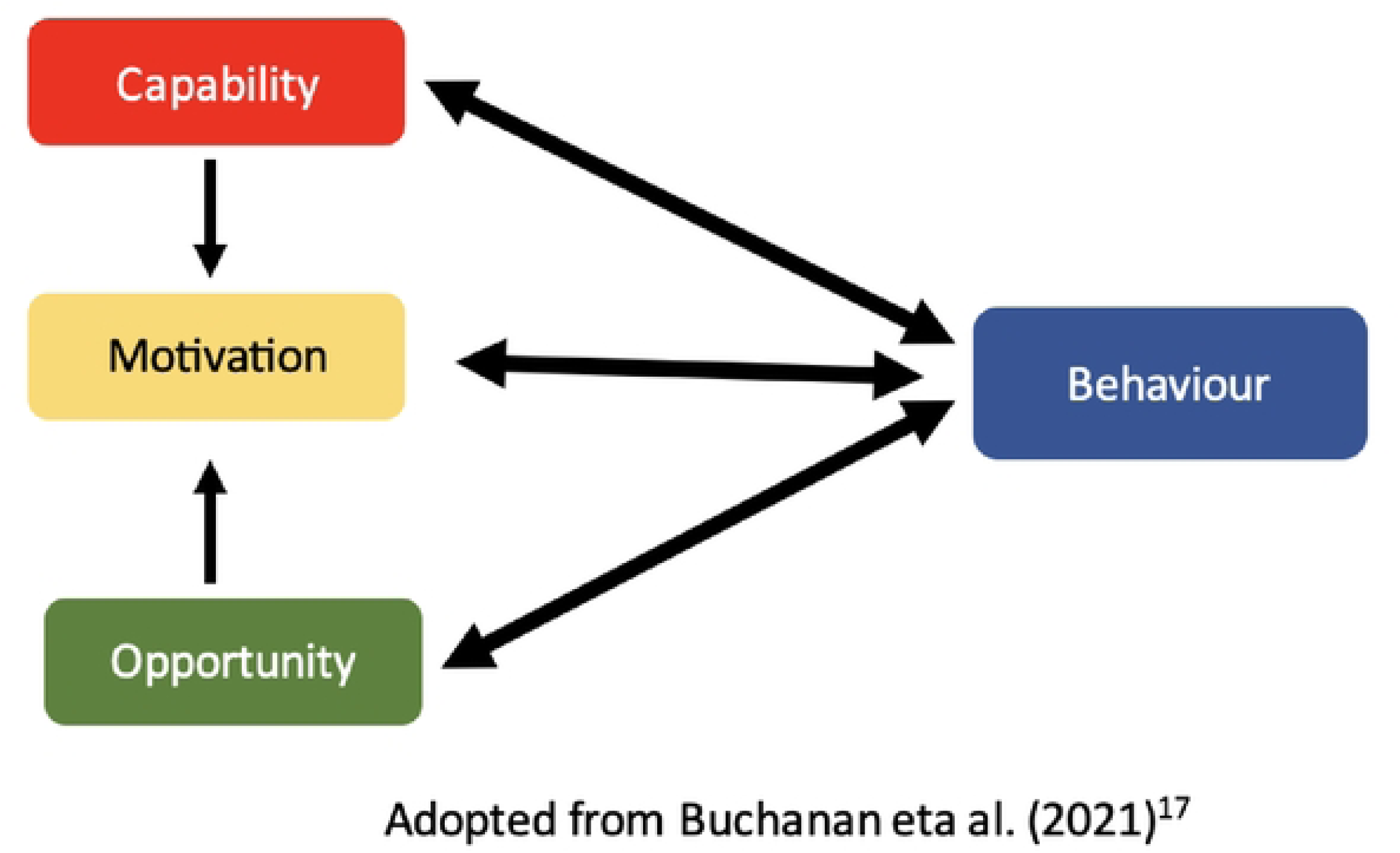
The COM-B Model

**Table 2:**
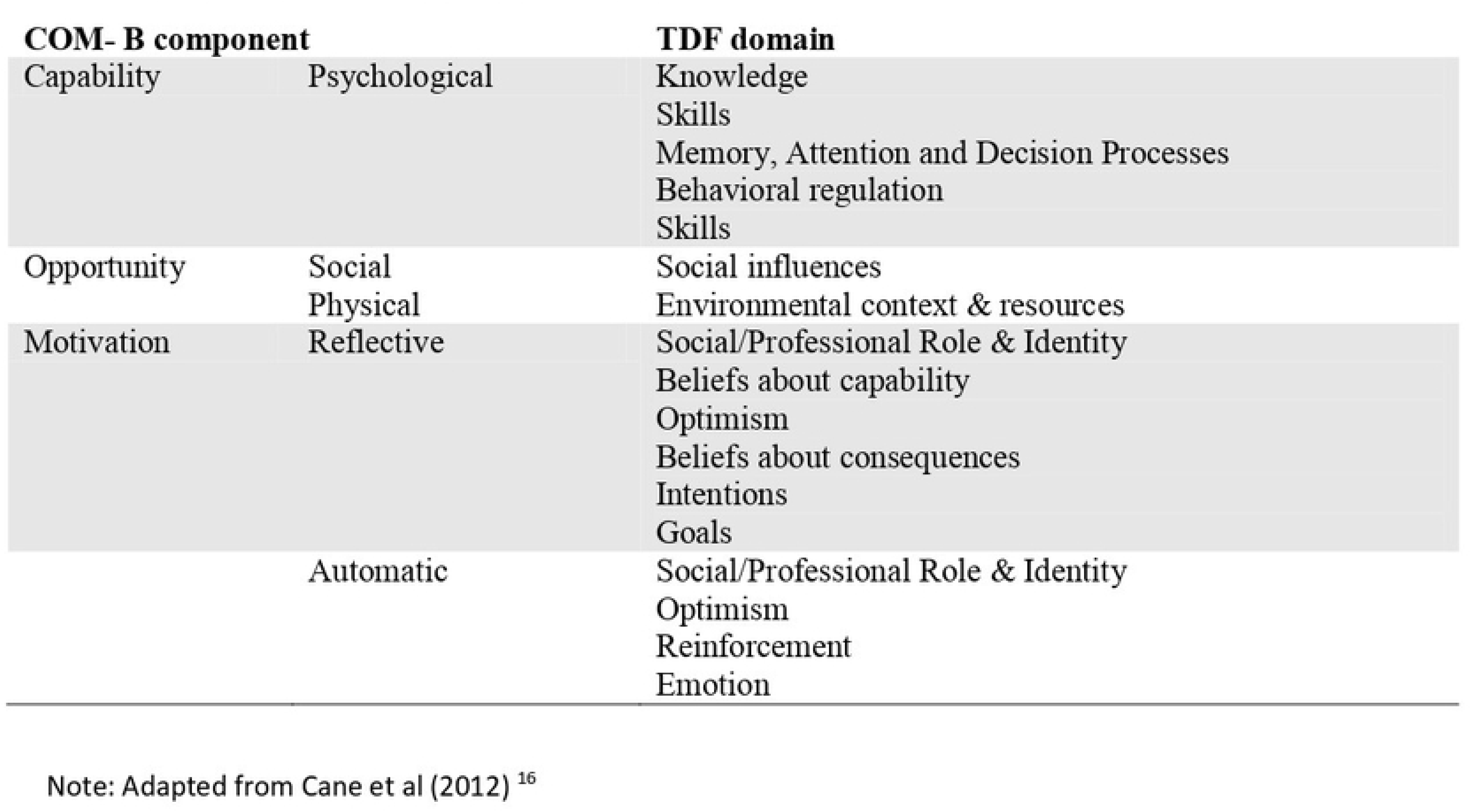
The COM-B Model and its relation to the TDF

Although the COM-B model and Theoretical Domains Framework (TDF) are effective tools for guiding health interventions, their use in CC screening research in the rural Kilimanjaro region is limited. This study aims to fill this gap by applying the COM-B and TDF models to explore facilitators and barriers to CC screening among women and healthcare providers in the rural Kilimanjaro region and ultimately supporting the design of theory-based interventions that can effectively drive behavior change and increase CC screening outcomes in the Kilimanjaro region(18).

## Materials and Methods

### Study Design

This study used an explorative qualitative formative design with an inductive-deductive framework (19).

#### Research Question

*What are the facilitators and barriers to cervical cancer screening among women and health care providers?*

#### Study settings

The study was conducted in the rural areas of the Kilimanjaro region, specifically in the Rombo and Moshi Rural districts, which are two of the seven districts in the region. The Kilimanjaro region has a total population of 1,861,934. Moshi Rural District counts 535,803 people, and Rombo District, 275,314 (Tanzania Census, 2022). These districts were chosen due to their higher prevalence of cervical cancer cases (36.4% in Moshi Rural and 15.2% in Rombo) and lower cervical cancer screening rates compared to other districts (38% in Moshi and 46% in Rombo) (KCMC Cancer Care Centre Registry database & Kilimanjaro Regional Cervical Cancer Screening Registry, 2021-2022). Rombo District has 2 hospitals, 5 health centers, and 38 dispensaries, while Moshi Rural District is home to 6 hospitals, 8 health centers, and 93 dispensaries. Cervical cancer screening services are available at the level of hospitals and health centers. Rombo District has 5 health facilities offering screening, while Moshi Rural District has 6 facilities providing the screening service. In total, the region has 32 facilities equipped to provide cervical cancer screening (Kilimanjaro Regional Cervical Cancer Screening Registry, 2021-2022).

#### Study population

The study population included screened women, irrespective of duration of screening and test results, and non-screened women living in the community under study. Non-screened women were included in this study to identify the factors hindering them from participating in cervical cancer screening. The inclusion criteria were women aged 30-50 years (age recommended for screening according to the Tanzania Cervical Cancer Prevention and Control Guideline, 2023), Swahili speakers, living in the study area, and those who consented to participate. Exclusion criteria included women on cervical cancer treatment.

Healthcare providers (nurses and doctors) providing cervical cancer screening services were also included from both private and public screening facilities in Rombo and Moshi rural districts, regardless of years of experience and level of education.

#### Selection of Health facilities

Six cervical cancer screening facilities were purposively selected for the study based on a 1:2 ratio corresponding to the population sizes of their respective districts: Moshi Rural District (535,803 people) and Rombo District (275,314 people). This included 2 facilities from Rombo and 4 from Moshi Rural District. Among the 6 selected facilities, 3 were privately owned. The facilities selected from Moshi Rural District included Kibosho Hospital, Kilema Hospital, TPC Hospital, and Himo OPD Health Centre, while those from Rombo District included Huruma Hospital and Karume Health Centre. The privately owned facilities were Kibosho Hospital, Kilema Hospital, and Huruma Hospital. These facilities were purposefully selected based on their geographical location, which ensures a diverse and representative sample population. The cervical cancer screening services provided by these facilities include visual inspection with acetic acid (VIA), follow-ups (tracking and reminding women of follow-up appointments and referrals for advanced care when necessary), pre- and post-screening education and counseling, treatment of precancerous lesions, and referrals.

#### Selection of Participants

Purposive sampling was used to recruit screened participants from the register provided by the Tanzanian Ministry of Health (MoH) at the health facilities. The register included information on serial number, client name, address, phone number, age, age at first sexual contact, HIV status, date of last menstrual period, screening results, use of cryotherapy, and any referrals for Loop Electrosurgical Excision Procedure (LEEP) or other treatments (20). The phone numbers listed on the screening register were used to contact study participants. After verifying the inclusion criteria and providing verbal summary information about the project by phone, an invitation to participate in the study was extended.

Purposive sampling was employed to recruit non-screened women from the community, as this method was well-suited for reaching the specific target population. Unlike random sampling, purposive sampling effectively identified non-screened women, a key demographic that might otherwise have been overlooked (21). Four community health workers (two in each district) were identified and acted as points of contact for women in the community. To ensure wide representation, the study recruited one woman per household; when more than one woman in a household was eligible, the study recruited the oldest (30–50 years). The study recruited twelve healthcare providers (nurses and doctors) whose primary responsibility was cervical cancer screening, with two participants selected from each of the chosen health facilities.

### Data collection

The Focus Group Discussion (FGD) guide was adopted from a previous study (22) without modifications, while the individual interview guide was adopted from two studies (23) and (24) and modified to exclude irrelevant questions, including those about clinic managers. Both tools were based on the COM-B model (capability, opportunity, motivation) and the TDF domains, which include knowledge, skills, social influences, beliefs, environmental context, resources, and social/professional role. The guides were translated into Swahili and back-translated by two independent experts to ensure accuracy (21).

Data were collected from 13^th^ March to 1^st^ October 2024. In total, six FGDs were conducted, three from each district, which included a group of screened women, non-screened women, and a mixed group of screened and non-screened women to explore differing perspectives on facilitators and barriers to cervical cancer screening. Each group had 8-12 number of participants. The inclusion criteria of mixed groups aimed to encourage open dialogue and explore shared and differing experiences between screened and non-screened women, providing comprehensive insights into screening behaviors. Skilled facilitation ensured a safe, inclusive environment that supported mutual understanding and open participation. The discussions were moderated by the lead author, with the support of two trained assistants (a female nurse and a male socio-scientist). FGDs were conducted in accessible and comfortable venues and lasted up to 75 minutes. Information collected included participants’ cervical cancer and screening histories, screening test experiences, information-seeking behaviors, social support, and suggestions for improvement.

Semi-structured in-depth interviews (IDIs) with healthcare providers were conducted at their workplaces and lasted up to 40 minutes. Data saturation was reached at 10 interviews. However, all 12 participants were interviewed to confirm that no new information emerged. Information collected from the individual interviews included providers’ knowledge, role, perceptions, emotional experiences, peer dynamics, and the barriers and facilitators involved in supporting behavior change.

FGDs and IDIs were audio-recorded by a digital device, and notes were taken. Informed consent was obtained from all participants. Each participant completed a socio-demographic information form, which took a maximum of five minutes to complete. Confidentiality was maintained through secure data handling practices.

#### Data management and synthesis

All FGDs and IDIs were audio-recorded, transcribed verbatim, and translated into English by VK and RM. Thematic analysis with an inductive-deductive approach was used (19). The consolidated criteria for reporting qualitative studies (COREQ) checklist was employed (25).

The transcripts were reviewed for accuracy and consistency with the audio recordings. The analysis included familiarization with the data, construction of a thematic framework based on the TDF domains, indexing, sorting data, and interpretation of data. An inductive-deductive coding framework was employed to avoid complications in the operationalization of the TDF domains (26). Using an inductive approach, two researchers (RM, VK) generated themes for similar data clusters, and definitions for these themes were developed. For the deductive element, themes were categorized into domains based on a TDF-based coding manual, which provided clear guidelines for how the inductively generated themes would be categorized within the TDF framework.

Two researchers (RM, VK) independently read and coded two transcripts. Recurrent themes and coding were compared and discussed to find differences for consensus. Once the coding framework was finalized, the remaining interviews were coded. Each theme was then deductively mapped to the TDF domains and subsequently to the COM-B model. The TDF framework identified key domains such as knowledge, beliefs, and social influences, guiding the development of study tools, while the COM-B model was used to design questions that assess capability, opportunity, and motivation, focusing on factors influencing screening participation. Themes were mapped to the three COM-B components and their subthemes: Capability (psychological), Motivation (automatic, reflective), and Opportunity (social, physical), as well as the eight TDF domains: Knowledge, Skills, Social Influences, Emotion, Environmental Context and Resources, Social/Professional Role and Identity, Beliefs about Consequences, and Beliefs about Capabilities, and were analyzed separately due to differences in the interview guides and the units of analysis. The findings from the analyses were combined during the write-up phase (27). Common verbatim debates related to facilitators and barriers were selected and included in the analysis to illustrate the findings. .

#### Trustworthiness of the study data

The criteria for data saturation—credibility, dependability, transferability, and confirmability were implemented to reduce the likelihood of bias influencing the researchers’ interpretations during data analysis. These criteria are strongly advocated in qualitative research to enhance the reliability of study outcomes(21,28). Credibility was strengthened by employing established research methodologies and collecting data directly from women and healthcare providers providing cervical cancer screening. Trustworthiness was upheld by selecting reliable participants and ensuring that the interviewer, who had no prior relationship with the participants, strictly followed the interview protocol. Additionally, content analysis guided the interpretation process, and two independent researchers reviewed the data separately. Their diverse perspectives promoted transparency and supported the study’s dependability and confirmability. Transferability was improved through detailed descriptions of the data collectors, data collection procedures, interview duration, and analysis methods. These details enable the findings to be applicable in other contexts (21,28).

#### Ethical Considerations

The study was conducted as per the Declaration of Helsinki (World Medical Association, 2013) and was approved by the KCMC University Research and Ethics Review Committee (Certificate No. 2642) and the National Institute of Medical Research (NIMR) in Tanzania (Certificate No. NIMR/HQ/R.8a/Vol. IX/4694). Verbal consent was initially obtained during the phone call, followed by written consent on the day of participation, with anonymity ensured using identification numbers. Nurses at study sites acted as key contacts for participant coordination. To reduce selection bias, researchers collaborated with nurses using predefined criteria to identify eligible participants from clinic registers; thereafter, the nurses contacted only those selected. Interviews were scheduled at participants’ convenient times to minimize interfering with their responsibilities.

## Results

### Characteristics of the study participants

Six FGDs involving 54 women were conducted, with 26 women screened and 28 not screened for cervical cancer. Women aged above 35 were 19 and 12 for screened and non-screened women, respectively. Additionally, 22 screened and 27 non-screened women had primary or secondary education, and nearly all participants were self-employed. IDIs were conducted among 12 health providers, 9 of whom were female. Four of the 12 participants had more than 10 years of experience in providing cervical cancer screening. Nearly all participants (10) had a college or university education, and almost half of the health providers or their spouses had not been screened for cervical cancer. Detailed information about the participants is shown in **Table 3**.

**Table 3:**
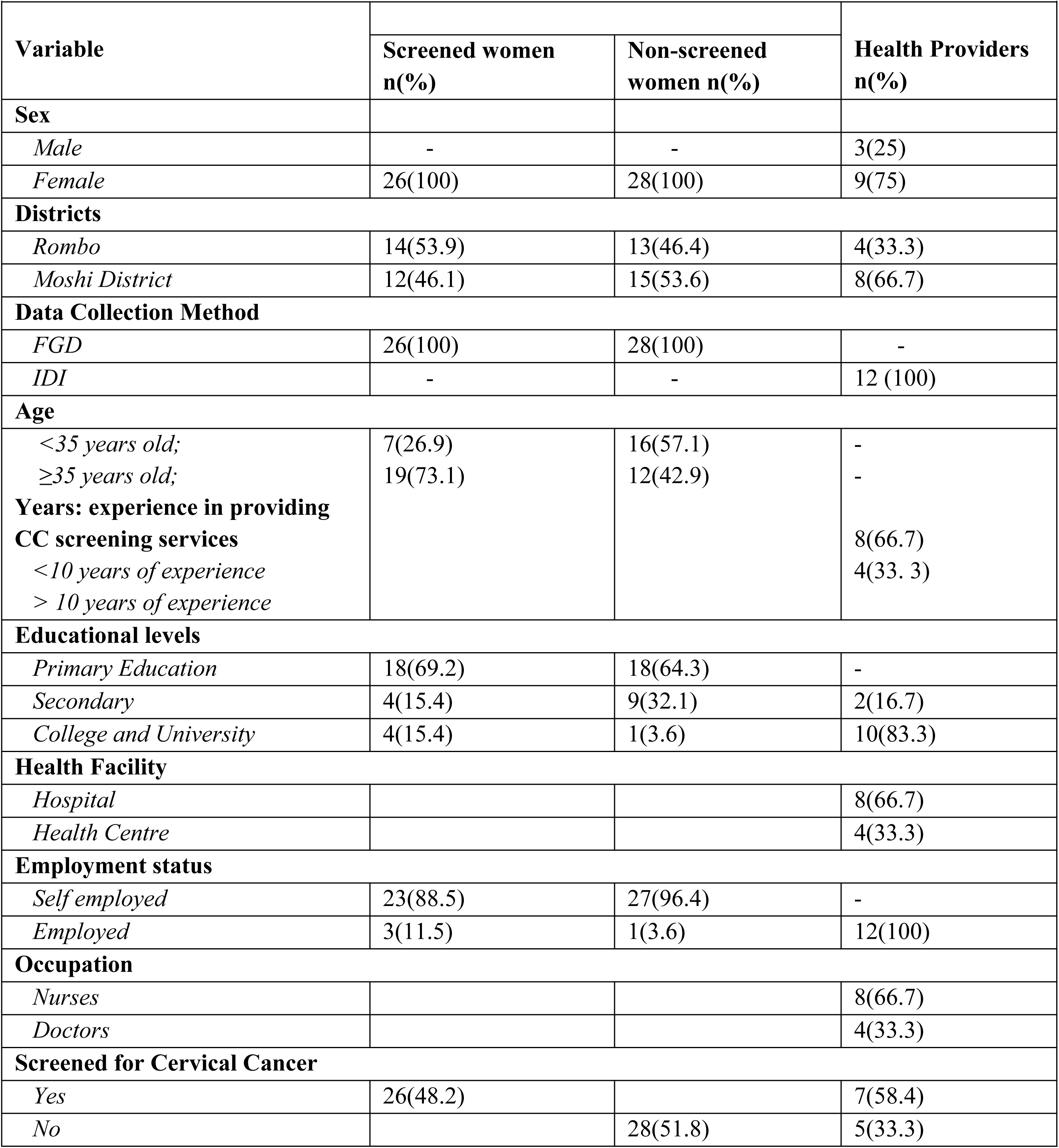
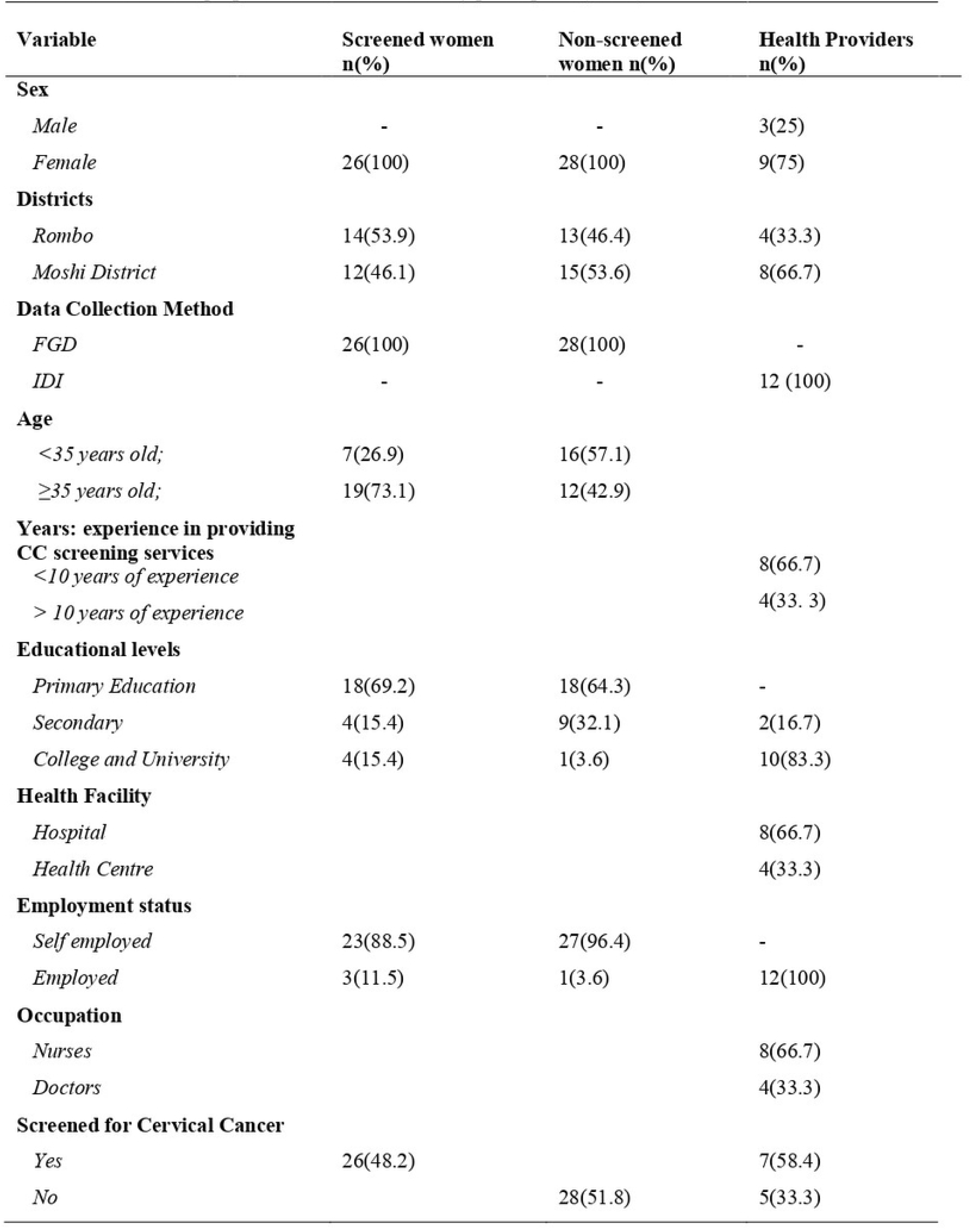
**Socio-demographic** characteristics of study participants (n= 66).

#### Summary of main themes mapped from TDF domains and COM-B constructs

Five main themes emerged from this study: 1. Knowledge of cervical cancer and screening, 2. Power of social Influence, 3. Emotional and structural influences on cervical cancer screening, and 4. Enhancing cervical cancer screening uptake. The results are presented for each main theme, corresponding to the COM-B constructs and TDF domain(s), featuring illustrative quotes throughout the main text.

#### Knowledge of cervical cancer and screening

This theme highlights a knowledge gap regarding cervical cancer between screened and non-screened women. Non-screened women only recognized the name of the disease but lacked knowledge of its meaning, causes, symptoms, and risk factors. Non-screened women did not know that cervical cancer is a serious health threat with a high mortality rate; they believed that screening was only necessary if symptoms or pain were present, not realizing that the disease can progress silently in its early stages. One of the non-screened participants reported that, “*Many people don’t know what cervical cancer is. They just hear the name, but if you ask them to explain, they won’t be able. There is very little knowledge, and that is a big problem” (FGD5-P2)*.

While the majority of screened participants’ demonstrated greater knowledge of cervical cancer and recognized it as a serious disease, which served as a motivation for undergoing screening. A screened woman explained*, “Cervical cancer develops without noticeable symptoms, making you feel healthy even if it’s progressing. Over time, it may reach an advanced stage, and only then you will start experiencing the pain. By the time you seek medical help, the condition may have become severe, and you could be in stage 4”* (FGD 2-P1). However, nearly all of the screened women had limited knowledge of the causes of cervical cancer.

Non-screened participants also showed limited knowledge of cervical cancer screening, including the HPV self-sampling tests. This was often attributed to insufficient education and a lack of accessible information. Several expressed a desire for more comprehensive education on the benefits of screening and the risks associated with not being screened: “*I wish to know the benefits of screening or the risks of not screening” (FGD5-P2)*. Furthermore, the study revealed confusion about which women are most at risk of developing cervical cancer and highlighted differing perceptions of screening between screened and non-screened participants. One non-screened participant said, “I *have never been screened because I have never had any problems” (FGD4-P9)*. The study found that healthcare providers were knowledgeable about cervical cancer screening, risk factors, eligibility, treatment, and referral procedures, which they gained through formal and on-the-job training. While expected, confirming their knowledge ensures accurate screening services and highlights potential practice inconsistencies. Healthcare providers recommended continuous education and refresher courses to enhance their skills.

There was a clear age-related difference, with most screened women being over 35 years old and most non-screened women under 35. This may be due to older women having more knowledge on cervical cancer, greater access to information, and more frequent contact with healthcare services. Parity could also play a role, as women with more children are more likely to use maternal health services where screening is promoted.

#### Power of Social Influence

This theme refers to the impact of social relationships and community dynamics in influencing women’s decisions to undergo cervical cancer screening. The theme is guided by the TDF domain of social influences and the COM-B model of social opportunity. We found that support from healthcare professionals, friends, family, religious leaders, and community leaders influenced women’s decisions to undergo cervical cancer screening. Screened participants reported that awareness campaigns at the community level have motivated them to seek screening; non-screened participants were still affected by misinformation and discouragement, suggesting that these campaigns did not effectively reach all community members.

One of the non-screened participants argued that, *“when we are in our groups, there are rumors here and there, like, hey, don’t dare go for screening; if they touch it, you will die”* (FGD5-P9). The study also highlighted the crucial role of healthcare providers (HCPs) in supporting women through education, addressing misconceptions, and offering assistance, such as follow-up calls and transportation. Participants acknowledged that HCPs’ involvement, along with community awareness campaigns and seminars, helps create social opportunities to spread information and motivate more women to participate in screening, thus contributing to family and community well-being. *“Mass screenings are helpful, so we should continue educating and advertising, perhaps in churches, mosques, and markets”* (IDI 01).

#### Emotional and structural influences on cervical cancer screening

This theme refers to the psychological and systemic factors that affect women’s willingness and ability to participate in cervical cancer screening. The theme mapped to the TDF domains of *Emotions* and *Environmental Context and resources,* and the COM-B components of *Automatic Motivation* and *physical opportunity,* explored both emotional and structural influences on cervical cancer screening.

Under the domain of *Emotions* and *Automatic Motivation*, both screened and non-screened women expressed fears related to pain, discomfort, and the possibility of receiving positive results. Screened women attributed their anxieties to personal experiences with the screening procedure, whereas community narratives and misconceptions influenced non-screened women. Despite these concerns, some screened women felt confident in coping with the procedure, recognizing the temporary nature of the pain and the seriousness of the disease. They also encouraged others to get screened: *“I understand this disease (meaning ‘cervical cancer’) is dangerous, and the pain experienced from speculum insertion is temporary” (FGD1-P3)*.

Screened participants highlighted relaxation techniques that helped manage stress, emphasizing the importance of psychological preparation and support. *“Learning about the screening process and having supportive guidance has made me more skilled in coping with the procedure. I now feel more confident in managing the screening experience.” (FGD3-P9)* HCPs found fulfillment when women accepted screening but noted that persistent myths and misconceptions continued to act as barriers to acceptance. Nevertheless, they remained committed to educating and motivating women, even when encouraging screening in the absence of symptoms proved challenging. *“I feel very motivated when I see that my counseling helps women take charge of their health.” (IDI 03)*

From the perspective of *environmental context and resources* and *physical opportunity*, several structural and systemic barriers were identified. Although screened women generally viewed access to services positively, they emphasized the need for ongoing education and greater awareness. Reported barriers included forgetfulness, discomfort with the speculum, limited availability of screening facilities, particularly in rural areas, and a shortage of trained healthcare personnel. These limitations hindered follow-up care and strained providers’ ability to offer adequate support and education. *“One of the biggest challenges is the shortage of healthcare providers. If we had more trained personnel, we could screen more women and provide better follow-up care.” (IDI 11)*.

#### Enhancing Cervical Cancer Screening Uptake

This theme refers to strategies, interventions, and factors to increase the number of women participating in cervical cancer screening. Theoretically, mapped to the TDF domain of Environmental Context and Resources, Social/Professional Role and Identity, Beliefs about Consequences, and Beliefs about Capabilities, and for the COM-B model—Reflective Motivation, highlighted improving cervical cancer screening uptake by making screening services more accessible in rural areas and promoting awareness through various channels. One of the screened participants responded that *“disseminating information or raising awareness within the community should not rely on just one method; a combination of methods is needed to reach the community effectively, such as through churches, mosques, organizing meetings in villages, broadcasting through media outlets, and using fliers for awareness”* (FGD2-R8).

Participants suggested using community meetings, religious gatherings, media, and fliers to spread information. While HCPs highlighted the challenge of reaching women outside hospitals, they recommended outreach in villages and markets to increase awareness and participation. “*Going to the villages to provide education on cervical cancer because many community members are unaware of these services and might never come to the hospital on their own” (IDI 06).* They stressed the importance of involving men in the process, as some husbands may discourage their wives from getting screened. Additionally, bringing screening services closer to communities, offering tests during sensitization campaigns, and providing clear results and treatment information were recommended to improve accessibility and encourage more women to participate.

## Discussion

Our study aimed to explore the facilitators and barriers to cervical cancer screening among women and healthcare providers. Four key themes emerged from the analysis. The findings revealed that major barriers included limited knowledge, misconceptions, and emotional fears related to pain and diagnostic outcomes. In contrast, key facilitators included access to accurate information, supportive interpersonal relationships, and active community engagement.

### Knowledge of cervical cancer and screening

We found that non-screened women in the rural Kilimanjaro region had limited knowledge about cervical cancer, its causes, and risk factors, often believing that screening was only necessary when symptoms or pain were present. Contributing factors included insufficient education, inadequate communication from healthcare providers, and widespread misconceptions. These findings are similar with studies conducted in Kenya [2022] (7), sub-Saharan Africa [2020] (10), Tanzania (2021] (20) and South Africa [2017] (29), which also identified limited knowledge about cervical cancer and deficiencies in provider’s education as significant barriers. Although screened women understood the seriousness of cervical cancer, their knowledge of its cause was limited, possibly due to health education focusing more on the screening process than on HPV. This gap may be exacerbated by shortages of healthcare providers. In contrast, a study conducted in Nigeria reported limited knowledge of HPV (30). Furthermore, non-screened women demonstrated limited knowledge of cervical cancer screening. This knowledge gaps among non-screened women increase their risks, highlighting the need for targeted education to support informed screening choices. Engaging community leaders and community health workers can boost screening rates and encourage behavior change. The knowledge difference between screened and non-screened women is likely the result of a combination of health system exposure (8), and social influence. These findings emphasize the importance of using the screening process as a platform for education, awareness, and expanding outreach efforts to reach women who do not typically access health services.

### Power of social influence

The present finding highlights the power of social support on informational (sharing accurate details about cervical cancer, screening, and its benefits), emotional (encouragement and reassurance), and practical roles (e.g., assistance with logistics like transportation) in facilitating cervical cancer screening uptake. Support from healthcare providers, family, friends, and religious leaders significantly influenced women’s screening decisions. Screened women reported that encouragement from family and personal interactions with healthcare professionals positively shaped their attitudes toward cervical cancer screening(31). In contrast, non-screened women often encountered negative social influences, including misinformation and discouragement, which impeded their engagement with screening services. Additionally, a disparity was observed between positive social influences and actual access to screening opportunities, particularly among women in underserved areas (22). These highlight the need for more inclusive and targeted approaches to ensure that no subgroup is left behind. Intervention should integrate education, emotional support, and community-based outreach. In a study conducted in Zimbabwe [2017] (12) further emphasized the importance of educating women, particularly those from traditional churches, to encourage cervical cancer screening. Enhancing social opportunities through targeted communication strategies and environmental planning may also increase cervical cancer screening participation, particularly among women facing social and logistical barriers (32).

### Emotional and structural influences on cervical cancer screening

This study highlights the combined impact of emotional and structural factors on cervical cancer screening, framed through the TDF and COM-B models. Women experienced emotional barriers, particularly fears of pain, diagnosis, and discomfort associated with the speculum (33). The origins of these fears varied, stemming from either personal experiences or community-driven misconceptions. In contrast, studies by Logan et al. [2011] (34) and Bateman et al. [2019](35) found that women frequently held negative perceptions of cervical cancer screening, often reporting feelings of fear and embarrassment.

Some screened women developed coping strategies, which boosted their confidence and willingness to encourage others to participate in screening. These results demonstrate the need for psychological support, education, and training to reduce emotional barriers and enhance women’s confidence. These results are similar to a study done in South Africa that supports community education programs and mass media campaigns to spread information about cervical cancer and address negative community perceptions (29)

Healthcare providers played a key role in motivating women but faced challenges in promoting screening among asymptomatic individuals, particularly outside clinical settings. Addressing these challenges requires broader community outreach to strengthen both psychological capability and practical opportunity for cervical cancer screening participation. In contrast, a 2017 study conducted in Kenya reported that women who underwent screening did so based on recommendations from their healthcare providers (33).

Structurally, the study highlights speculum discomfort, limited service availability, and shortages of healthcare providers, especially in rural areas, as significant barriers. These findings emphasize the need for theory-informed interventions that address both individual and systemic obstacles to improve screening uptake, including education, peer support, and service delivery enhancements.

#### Enhancing Cervical Cancer Screening Uptake

The present study emphasized the need for improved access to education and services. Utilizing diverse outreach platforms where most women are found, such as religious gatherings, markets, community meetings, women’s groups, and social media, can raise awareness and improve knowledge, especially in rural communities (36,37). Participants indicated that understanding cervical cancer would motivate screening and recommended mobile clinics and the use of community health workers to address distance barriers (24). Involving community leaders in advocacy enhances trust, reduces stigma, and promotes culturally accepted health initiatives. This is supported by Fang et al. 2019 (38), who reported that community-based educational programs can enhance awareness of cervical cancer and screening, foster positive attitudes towards the benefits of screening, and help reduce perceived barriers to accessing it. Additionally, engaging men in cervical cancer screening promotion is crucial, as their support influences women’s health decisions; however, the majority of men reported having limited information on cervical cancer screening (39). Findings from this present study also supported the HPV self-sampling test, highlighting its comfort and potential to increase screening if made affordable and accessible. A comprehensive approach that combines education, community involvement, service accessibility, male engagement, and HPV self-sampling testing was reported to enhance screening uptake, reduce mortality, and improve health equity and outcomes in rural populations.

### Strengths and limitations

This study demonstrated several qualitative strengths that contributed to the trustworthiness and rigor of its findings. It employed the four widely recognized criteria of qualitative trustworthiness: credibility, dependability, transferability, and confirmability (Lincoln & Guba, 1985; Polit & Beck, 2017). Data were collected directly from women and healthcare providers actively involved in cervical cancer screening, ensuring relevance and capturing firsthand experiences. The interviewer maintained neutrality, having no prior relationship with participants and strictly adhering to the interview protocol to minimize potential bias. Additionally, content analysis was used to guide interpretation, and two independent researchers reviewed the data separately, thereby strengthening dependability and reducing individual bias. Detailed descriptions of the data collection process, interview procedures, and analytical methods further enhanced transferability, allowing the findings to be considered in other similar contexts.

The study faced several limitations. The participant scope was limited to women and healthcare providers, excluding administrative personnel, but they are planned to be included in the stakeholders meeting to discuss the identified gaps and support planning for the intervention. Moreover, the sample included a disproportionate number of nurses compared to doctors, reflecting the staffing structure of the clinics but potentially skewing the findings toward a nursing viewpoint. Despite measures to reduce bias, the researchers’ interpretations could still have been influenced by their assumptions. Although participant validation was employed to enhance credibility, this process may have caused participants to modify or withhold certain information. Lastly, while multiple coders helped improve reliability, the consensus approach may have limited the exploration of alternative interpretations of the data.

### Conclusions

Cervical cancer screening uptake can be improved by strengthening facilitators such as social support and addressing existing barriers, including limited knowledge, misconceptions, fear of pain and diagnosis, and service inaccessibility. The findings highlight the need for theory-based interventions that incorporate education, community engagement, and health system improvements to increase screening uptake in rural communities.

## Data Availability

All data used in this manuscript will be available

## Acknowledgments

The authors gratefully acknowledge the study participants, screening sites, district religious heads, and district executive officers from Rombo and Moshi rural districts.

## Supporting information

S1 FGD Analysis Table

S2 Analysis Table IDI

